# Effects of a Water, Sanitation, and Hygiene Program on Diarrhea and Child Growth in the Democratic Republic of the Congo: A Cluster-Randomized Controlled Trial of the Preventative-Intervention-for-Cholera-for-7-Days (PICHA7) Program

**DOI:** 10.1101/2024.12.16.24318942

**Authors:** Christine Marie George, Presence Sanvura, Jean-Claude Bisimwa, Kelly Endres, Alves Namunesha, Willy Felicien, Blessing Muderhwa Banywesize, Camille Williams, Jamie Perin, David A Sack, Raissa Boroto, Gisèle Nsimire, Feza Rugusha, Freddy Endeleya, Pacifique Kitumaini, Claude Lunyelunye, Emmanuel Buhendwa, Pascal Kitumaini Bujiriri, Jessy Tumusifu, Brigitte Munyerenkana, Laura E. Caulfield, Justin Bengehya, Ghislain Maheshe, Cirhuza Cikomola, Alain Mwishingo, Lucien Bisimwa

**Affiliations:** Department of International Health, Program of Global Disease Epidemiology and Control. Johns Hopkins Bloomberg School of Public Health, Baltimore, MD 21205, USA; Center for Tropical Diseases & Global Health, Université Catholique de Bukavu, Bukavu B.P 265, Democratic Republic of the Congo; Department of International Health, Program in Human Nutrition, Johns Hopkins Bloomberg School of Public Health, Baltimore, MD 21205, USA; Bureau de l’Information Sanitaire, Surveillance Epidémiologique et Recherche Scientifique, Division Provinciale de la Santé Sud Kivu, Ministère de la Santé, Bukavu B.P 265, Democratic Republic of the Congo; Faculty of Medicine, Catholic University of Bukavu, Bukavu B.P 265, Democratic Republic of the Congo

**Keywords:** diarrhea, mobile health, child growth, randomized controlled trial

## Abstract

**Background:** We assessed whether the Preventative-Intervention-for-Cholera-for-7-Days (PICHA7) program reduced diarrhea and improved child growth in the Democratic Republic of the Congo (DRC).

**Methods:** The PICHA7 cluster-randomized controlled trial enrolled diarrhea patient households in urban Bukavu, DRC. Households were randomized into two arms: single in-person visit for the DRC government’s diarrhea patient standard message on oral rehydration solution use and a basic WASH message (standard arm); or this standard message and the PICHA7 program with quarterly in-person visits and weekly voice and text mobile health messages (mHealth) (PICHA7 arm). The primary outcome was diarrhea in the past two weeks assessed monthly for 12 months. Secondary outcomes were diarrhea with rice water stool, healthcare facility visits for diarrhea, stunting, underweight, and wasting over 12 months. Generalized estimating equations were used for regression models to account for clustering at the individual and household level.

**Results:** Between December 2021 and December 2022, 2334 participants in 359 households were randomly allocated to two arms: 1138 standard arm and 1196 PICHA7 arm. Diarrhea prevalence during the 12 month surveillance period was significantly lower among PICHA7 arm participants (Prevalence Ratio: 0.39 (95% Confidence Interval (CI): 0.32, 0.48). PICHA7 arm participants had lower odds of diarrhea with rice water stool (Odds Ratio (OR): 0.48 (95% CI: 0.27, 0.86)), and lower odds of visiting a healthcare facility for diarrhea during the 12 month surveillance period (OR: 0.44 (95% CI: 0.25, 0.77)). PICHA7 arm children 0-4 were significantly less likely to be stunted (52% vs. 63% standard arm) (OR: 0.65 (95% CI: 0.43, 0.98)) at the 12 month follow-up. All WASH components had high adherence.

**Conclusion:** The PICHA7 program which combines mHealth with quarterly in-person visits lowered diarrhea prevalence and stunting in the DRC.

**Clinical Trials Registration:** NCT05166850.

**Key Points:** The PICHA7 program combines mHealth with in-person visits and was assessed with a randomized controlled trial. Compared to the standard arm, the PICHA7 program lowered diarrhea prevalence for all ages and reduced stunting in children 0-4 years in the DRC.

## Introduction

Globally there are an estimated 2.3 billion diarrhea episodes annually.^1^ The highest burden of diarrheal disease morbidity and mortality is among children 0-4 years in sub-Saharan Africa.^1,2^ Frequent enteric infections among young children can result in intestinal inflammation leading to reduced nutrient absorption which has been associated with impaired linear growth in young children.^3^ In 2022, 148 million children 0-4 years were estimated to be stunted globally.^4^ Stunting has been associated with poor cognitive outcomes and a higher risk of mortality and chronic diseases later in life.^5,6^ Effective water, sanitation, and hygiene (WASH) interventions are urgently needed to reduce diarrheal disease outbreaks and improve child growth globally.

Worldwide there are an estimated 2.9 million cholera cases annually.^7^ The Democratic Republic of the Congo (DRC) has one of the highest rates of cholera in Africa.^8^ Individuals living in close proximity to cholera patients are at an increased risk of subsequent cholera infections.^9^ A recent study in the DRC found individuals living within 30 meters of a cholera patient were at 20 times higher risk of developing cholera during the 7-day period after patient hospitalization, with studies from other settings showing elevated risk continuing for one month.^9,10^

In an effort to reduce diarrheal diseases among the household members of diarrhea patients, our research group developed the Preventative-Intervention-for-Cholera-for-7-Days (PICHA7) program in eastern DRC through 18 months of community-centered formative research.^11^ “Picha” means picture in Swahili for the pictorial WASH module delivered as part of the program and “7” indicates the 7-day high risk period after the diarrhea patient is admitted to a healthcare facility for subsequent diarrheal disease for diarrhea patient household members. The PICHA7 program is healthcare facility initiated through an initial healthcare facility visit delivered by a health promoter to diarrhea patients and their household members. This program is then reinforced through the PICHA7 mobile health program delivered through weekly voice and text messages and quarterly home visits during a 12-month period.

We investigated whether delivery of the PICHA7 program through weekly voice and text messages and quarterly in-person visits to diarrhea patient households could lower diarrhea prevalence, improve child growth, and increase handwashing with soap, water quality, and water treatment behaviors during the 12-month program period.

## Methods

### Study Design

The PICHA7 study was a two-arm cluster RCT conducted in urban Bukavu in South Kivu Province of eastern DRC from December 22, 2021 to December 20, 2023. An individual diarrhea patient household is defined as a cluster. Diarrhea patients were recruited from 115 public and private healthcare facilities in Bukavu City. This study compared a single in-person healthcare facility visit to deliver the DRC government’s standard message to diarrhea patients on oral rehydration solution use for dehydration and a basic 5-minute WASH message (standard arm) to this standard message plus the PICHA7 program. The PICHA7 intervention includes a single in-person healthcare facility visit to deliver a WASH module, a mHealth program with weekly voice, interactive voice response, and text messages sent to patient households, and quarterly home visits to reinforce WASH behaviors for 12-months. The Catholic University of Bukavu (7107) and the Johns Hopkins Bloomberg School of Public Health Institutional Review Board (9848) approved the study protocol.

### Participants

To be eligible diarrhea patients had to: 1) be admitted to a healthcare facility with three or more loose stools over a 24-hour period; 2) have no running water inside of their home (mostly slum areas); 3) provide a blood and a stool sample within 24-hours of enrollment (for etiology of diarrhea); 4) plan to reside in Bukavu for the next 12-months (assessed during screening prior to study enrollment); 5) have a child 0-4 years in household; and 6) have a least one working mobile phone in household. There was no age restriction for index diarrhea patients, all age groups were included if found to be eligible. Household members of the diarrhea patient were eligible for the trial if: 1) they shared the same cooking pot and resided in the same home with the diarrhea patient for the last three days; and 2) planned to reside with the diarrhea patient for the next 12-months (assessed during screening prior to study enrollment). Recruitment of diarrhea patients occurred daily from December 2021 to December 2022. All study participants or their guardians provided written informed consent.

### Randomization

Randomization of diarrhea patients to study arms (1:1) was performed using a random number generator. The study biostatistician (JP) assigned study randomization using R (version 3.3.0). JP was not involved in trial data collection. Randomization was performed after the baseline enrollment of the diarrhea patient. The study arm assignment was revealed to the study coordinator in a digital database by the study manager (ASP). Blinding was not possible because of visible intervention components. To minimize bias, we used separate teams for intervention and evaluation activities.

### Intervention Procedures

We developed the PICHA7 program through 18-months of community-centered formative research using a theory-informed approach guided by the Integrated Behavioral Model for Water, Sanitation and Hygiene (IBM-WASH).^11-13^ Formative research included semi-structured interviews and a pilot study of 518 participants. A detailed description of the intervention protocol is published elsewhere.^11^ The PICHA7 program targets five key behaviors: (1) preparing soapy water using water and detergent powder or water and small leftover bar soap pieces; (2) handwashing with soap or ash at food- and stool-related events; (3) safe drinking water storage in a water vessel with a lid and tap; (4) treating household drinking water using chlorine tablets; and (5) heating of household drinking water until it reaches a rolling boil after the 7-day high risk period. The PICHA7 program was initially designed to focus on cholera patients and their household members. However, based on the recommendation of the DRC Ministry of Health this WASH program was broadened to include diarrhea patients of all etiologies to allow the program to benefit millions more beneficiaries in DRC. The PICHA7 program is described in detail in Supplementary File 1.

### Outcomes

The primary outcome was self- or caregiver-reported diarrhea prevalence captured at monthly clinical surveillance visits during the 12-month study period (at a total of 12 timepoints). Diarrhea prevalence was defined as three or more loose stools in a 24-hour period in the past two weeks and was investigated among all age groups including children 0-4 and 0-1 years. Severe diarrhea (secondary outcomes assessed monthly which include all participants) was defined as participants visiting a healthcare facility for diarrhea treatment and participants with diarrhea with rice water stool in the past two weeks (key symptom of cholera^14^) at least once during study period. Child growth (secondary outcome) included the following measures of child growth for children 0-4 years: 1) stunting (height-for-age Z-scores (HAZ) <-2); 2) underweight (weight-for-age Z-scores (WAZ) Z <-2); and 3) wasting (weight-for-height (WHZ) Z-scores <-2). WASH related secondary outcomes were: 1) observed handwashing with soap at stool- and food-related events; 2) *Escherichia coli* (*E.coli*) in stored drinking water; and 3) free chlorine in stored drinking water <0.2 and <0.5 mg/L.

Research officers conducting evaluation activities received a 3-month formal training during the pilot study period prior to the RCT start. Diarrhea prevalence was assessed during monthly clinical surveillance visits. For children <5 years of age, research officers trained in standardized anthropometry measured each child’s weight and length (<2 years)/height (≥2 years) three times at baseline and at a 12-month follow-up.^15^ Seca infant length boards model 417 were used for children <2 years and Seca model 213 height boards for children 2-4 years. The average of these three measurements was taken at each timepoint. The window for anthropometric measurements at baseline was <48 hours of enrollment, and for the 12-month follow-up the anthropometric measurement was collected at ≥365 days of enrollment. Due to the COVID-19 pandemic and ongoing conflict at our study site a subset of children had baseline anthropometric measurements collected within the required window of 48 hours after enrollment. HAZ, WAZ, and WHZ were calculated according to the WHO child growth standards.^16^

Unannounced spot checks to prevent households preparing for the study team arrival were performed in a random subset of 100 households per study arm at Day 7 and 1, 3, 6, 9, and 12 months after enrollment to collect a household stored water sample to measure free chlorine and *E.coli*. The WHO guideline for drinking water quality of <1 colony forming units (CFU) /100 mL of *E.coli* in drinking water was used as the cutoff to define safe drinking water quality relative to microbial contamination.^17^ The WHO free chlorine guidelines of >0.2 and >0.5 mg/L for drinking water were used as cutoffs for a safe minimum drinking water level of free chlorine.^18^ Five-hour structured observations were conducted in a random subset of 50 households per study arm at Day 7 and 1, 3, 6, 9, and 12 months after enrollment. Handwashing with soap was recorded at the following key events promoted in PICHA7: 1) after using the toilet, 2) after cleaning a child’s anus, 3) after removing a child’s feces; 4) before eating; 5) before feeding a child; 6) before preparing food; and 7) before serving food.

### Statistical Analysis

The sample size calculation for diarrhea prevalence was based on a diarrhea prevalence of 10% for all ages using Global Burden of Disease Study^19^ estimates and 22% for children 0-4 years from the DRC Demographic Health Survey.^20^ Estimates assumed a 25% minimum detectable difference between study arms.^21^ The calculation assumed a type I error α of 0.05 and a power (1-β) of 0.80 and 6 individuals per household (based on PICHA7 pilot surveillance data). The sample size calculation indicated 250 diarrhea patient households per study arm (500 household total, 3000 participants), assuming a 23% loss to follow-up (based on the PICHA7 pilot), to detect a 2.5% change in diarrhea prevalence for all ages (10% standard arm vs. 7.5% in the intervention arm) and to detect 5.5% change in diarrhea prevalence for children 0-4 years (22% vs. 16.5%).

We analyzed participant data according to their randomized assignment (intention-to-treat), irrespective of their adherence to the assigned intervention. The statistical analysis plan implemented was the same as that for the Cholera Hospital Based Intervention for 7 Days (CHoBI7) RCT (NCT04008134), our RCT conducted in Bangladesh with the identical study design for evaluation activities.^22^ This statistical analysis plan was finalized prior to data analysis. Log binomial regression was performed to estimate the prevalence ratio (PR) for diarrheal disease using generalized estimating equations (GEE) to account for clustering at the individual (repeated surveillance visits) and household level and to approximate 95% confidence intervals (CI). Diarrhea prevalence was calculated using all monthly visits during the 12-month surveillance period and is defined as a 12-month diarrhea prevalence (multiple observations are included for each individual). Logistic regression was performed to estimate odds ratios (OR) for binary growth measures using GEE to account for household clustering. Models for diarrheal disease were adjusted for follow-up timepoint (timepoint for each observation was included in the model to adjust for seasonality) and models for child growth were adjusted for baseline growth measurements. For the growth analysis, we excluded observations with z-scores outside the biologically plausible ranges according to WHO recommendations.^16^ Logistic regression models were performed to compare intervention fidelity indicators by study arm using GEE to account for household clustering and approximate 95% CIs. Analyses were performed in SAS (version 9.4). The trial is registered at ClinicalTrials.gov (NCT05166850). Lost to follow-up was defined as individuals with no diarrhea prevalence data after baseline (individuals that did not contribute to the analysis for the primary study outcome).

## Results

Research officers identified 853 diarrhea patients with a child 0-4 years in their household during screening, 40% (337) were found to be ineligible. The most common reason for ineligibility was having a tap with running water in one’s home at 30% (101/337). Sixty-nine percent (359/516) of diarrhea patient households eligible for the study agreed to participate from 87 health facilities. Between December 2021 and December 2022, we randomly allocated 359 diarrhea patient households (2334 participants) to the standard and PICHA7 program arms (Figure 1). A map of the study area is shown in Supplementary File 2. Thirteen percent of participants (338) were lost to follow-up after baseline enrollment. When a sensitivity analysis was performed comparing participants lost to follow-up after baseline vs. those participants available during the study follow-up period there were no significant difference observed in baseline diarrhea prevalence. Nineteen participants died during the study period (6 adults and 13 children). Enrolled household baseline characteristics were similar across study arms (Table 1). Most study households had at least one person that could read and write (98%). Thirty-three percent of children 0-4 years (259/794) were index diarrhea patients in their households (35% in the standard arm and 30% in the PICHA7 arm).

**Figure 1.**
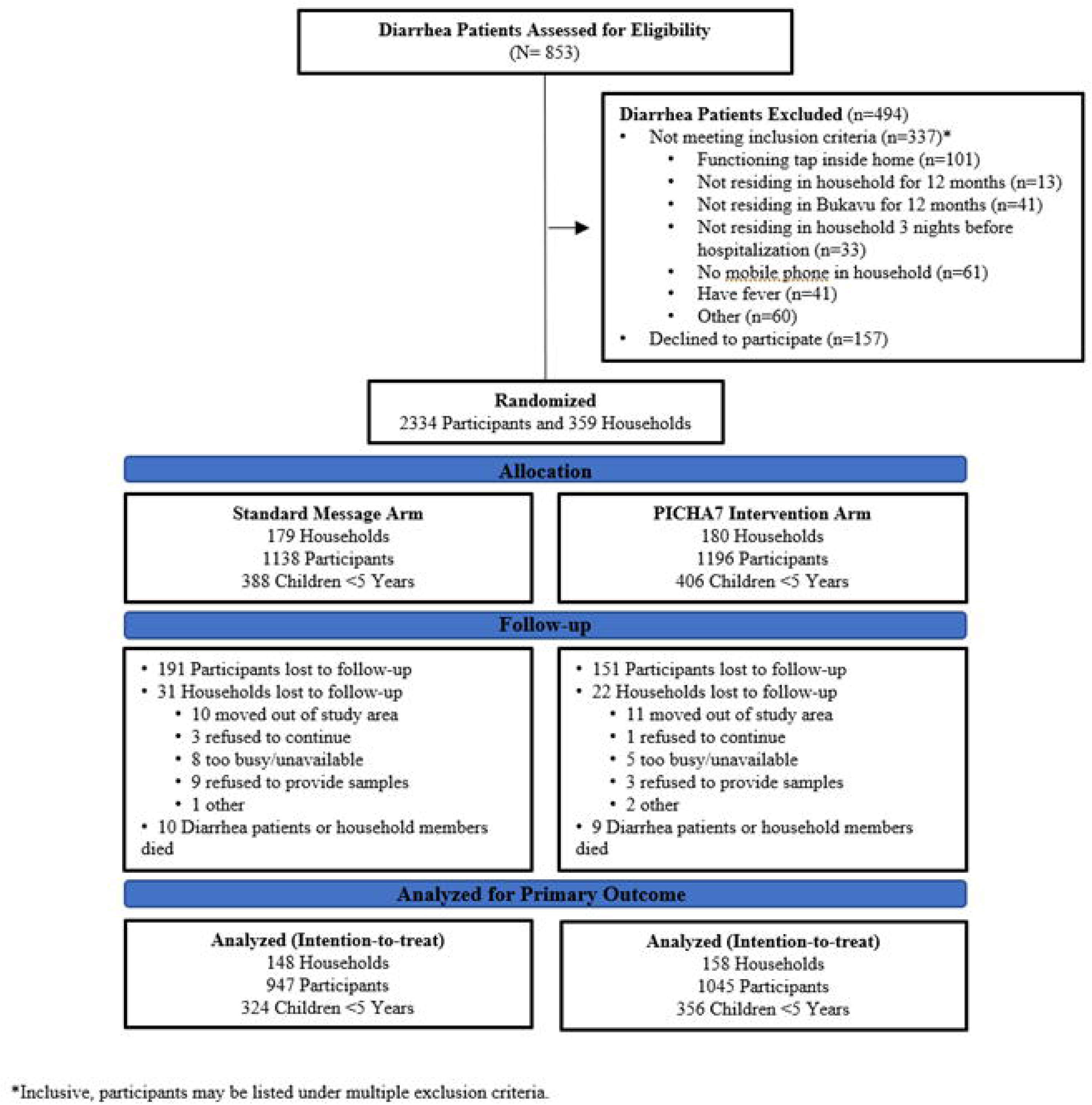
Trial profile and analysis populations for primary outcomes.

**Table 1.** Baseline Population Characteristics by Study Arm.

|  | Standard Message Arm |  |  | PICHIA7 Arm |  |  |
| --- | --- | --- | --- | --- | --- | --- |
|  | % | n | N | % | n | N |
| **Index Diarrhea Patient Households** |  |  | 179 |  |  | 180 |
| **All Study Participants** |  |  | 1138 |  |  | 1196 |
| **Household Members of Index Diarrhea Patients** | 84% | 957 | 1138 | 85% | 1016 | 1196 |
| **Children 0-4 Years** | 34% | 388 | 1138 | 34% | 406 | 1196 |
| **Baseline Participant Age** |  |  |  |  |  |  |
| Mean ± SD (Min - Max) (Years) | 13.2 ± 13.3 (0 - 70.7) |  |  | 13.6 ± 13.8 (0 - 78.5) |  |  |
| 0-1 Years | 19% | 212 | 1138 | 18% | 213 | 1196 |
| 2-4 Years | 15% | 176 | 1138 | 16% | 193 | 1196 |
| 5-17 Years | 38% | 435 | 1138 | 38% | 454 | 1196 |
| 18 Years or Greater | 28% | 315 | 1138 | 28% | 336 | 1196 |
| **Gender** |  |  |  |  |  |  |
| Female | 58% | 663 | 1138 | 59% | 709 | 1196 |
| **Household Size** |  |  |  |  |  |  |
| Mean ± SD (Min - Max) (Years) | 7.2 ± 2.6 (3 - 16) |  |  | 7.3 ± 2.7 (1 - 15) |  |  |
| **Household Roof Type** |  |  |  |  |  |  |
| Concrete | 6% | 10 | 169 | 5% | 8 | 171 |
| Other | 94% | 159 | 169 | 95% | 163 | 171 |
| **Household income** |  |  |  |  |  |  |
| Household income | 93% | 156 | 168 | 96% | 163 | 170 |
| No Household Income | 7% | 12 | 168 | 4% | 7 | 170 |
| **Refrigerator Ownership** | 4% | 6 | 169 | 4% | 6 | 171 |
| **Baseline Water Source** |  |  |  |  |  |  |
| < 1 CFU per 100 mL of *E.coli* | 65% | 64 | 99 | 61% | 57 | 93 |
| 1-10 CFU per 100 mL of *E.coli* | 11% | 11 | 99 | 15% | 14 | 93 |
| 10-100 CFU per 100 mL of *E.coli* | 11% | 11 | 99 | 11% | 10 | 93 |
| >100 CFU per 100 mL of *E.coli* | 13% | 13 | 99 | 13% | 12 | 93 |
| **At Least One Household Member Can Read and Write** | 98% | 165 | 168 | 98% | 168 | 171 |
| **Anthropometric Measurements for Children 0-4 Years** |  |  |  |  |  |  |
| Stunting at baseline (HAZ < -2) | 52% | 69 | 133 | 48% | 70 | 146 |
| HAZ Mean ± SD | -2.17 ± 1.68 |  |  | -1.93 ± 1.38 |  |  |
| Underweight at baseline (WAZ < -2) | 27% | 37 | 135 | 22% | 33 | 147 |
| WAZ Mean ± SD | -1.26 ± 1.49 |  |  | -1.01 ± 1.27 |  |  |
| Wasting at baseline (WHZ < -2) | 9% | 12 | 132 | 4% | 6 | 141 |
| WHZ Mean ± SD | -0.15 ± 1.66 |  |  | 0.09 ± 1.50 |  |  |

SD=Standard Deviation

Among all participants (all ages), 12-month diarrhea prevalence was significantly lower (diarrhea prevalence over the 12-month study period assessed through monthly visits) in the PICHA7 program arm (Prevalence Ratio (PR): 0.39 (95% Confidence Interval (CI): 0.32, 0.48) compared to the standard arm (Table 2). The impact on 12-month diarrhea prevalence was similar for children 0-4 years (PR: 0.38, 95% CI: 0.31, 0.46) and children 0-1 years (PR: 0.43, 95% CI: 0.35, 0.53)). Supplemental Table 1 includes analyses by age group and gender.

**Table 2.**
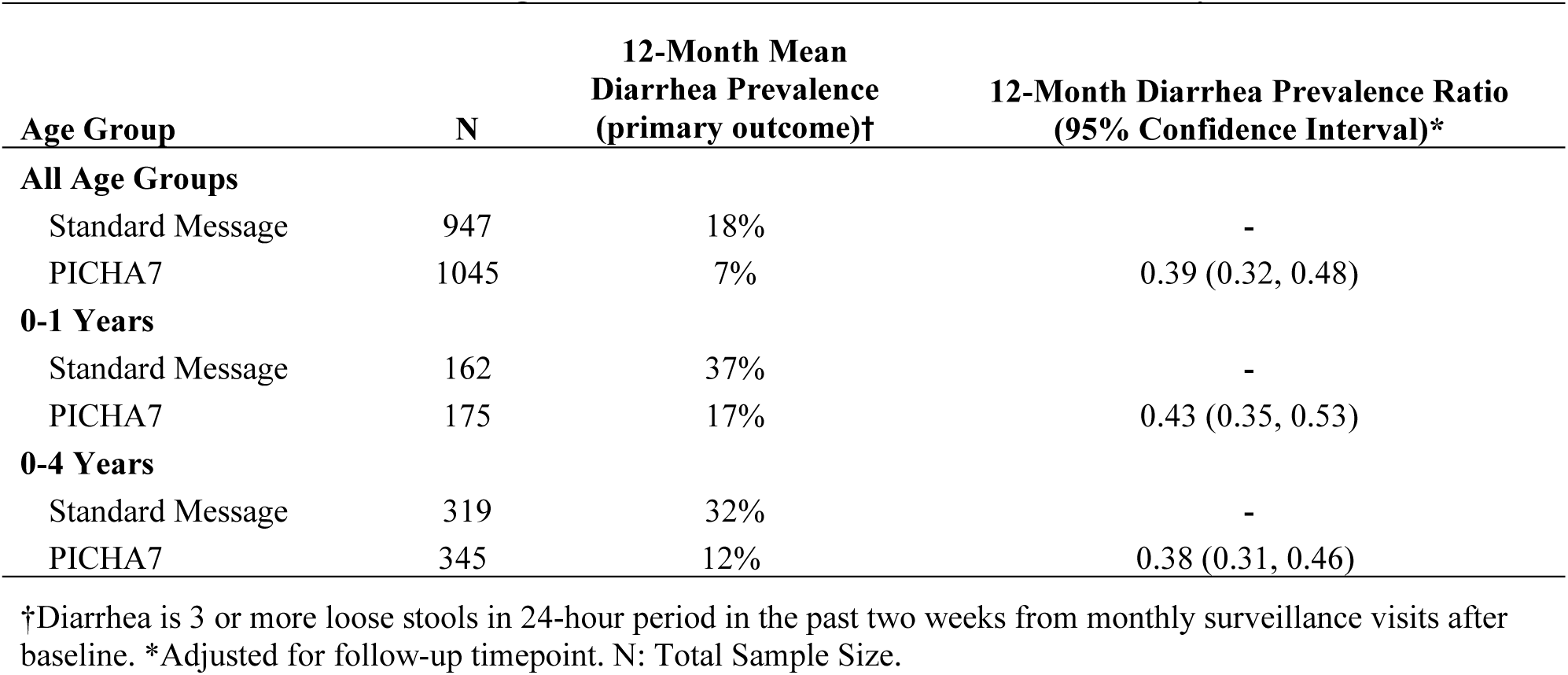
Effects of the PICHA7 Program on 12-Month Diarrhea Prevalence (Primary Outcome)

| **Age Group** | **N** | **12-Month Mean**<br>**Diarrhea Prevalence**<br>**(primary outcome)†** | **12-Month Diarrhea Prevalence Ratio**<br>**(95% Confidence Interval)\*** |
| --- | --- | --- | --- |
| **All Age Groups** |  |  |  |
| Standard Message | 947 | 18% | - |
| PICHA7 | 1045 | 7% | 0.39 (0.32, 0.48) |
| **0-1 Years** |  |  |  |
| Standard Message | 162 | 37% | - |
| PICHA7 | 175 | 17% | 0.43 (0.35, 0.53) |
| **0-4 Years** |  |  |  |
| Standard Message | 319 | 32% | - |
| PICHA7 | 345 | 12% | 0.38 (0.31, 0.46) |

†Diarrhea is 3 or more loose stools in 24-hour period in the past two weeks from monthly surveillance visits after baseline. \*Adjusted for follow-up timepoint. N: Total Sample Size.

Participants in the PICHA7 arm had a 52% lower odds of diarrhea with rice water stool during the 12-month surveillance period compared to the standard arm (all age groups) (OR: 0.48 (95% CI: 0.27, 0.86)) (4% vs. 2% (N=1992)). There was a 56% lower odds of participants visiting a healthcare facility for diarrhea during the 12-month surveillance period in the PICHA7 arm compared to the standard arm (all age groups) (OR: 0.44 (0.25, 0.77)) (5% vs. 2% (N=1981)).

Children 0-4 years were significantly less likely to be stunted at the 12-month follow-up in the PICHA7 arm compared with the standard arm (52% vs. 63%) in the unadjusted model (OR: 0.65 (95% CI: 0.43, 0.98)) and after adjustment for baseline growth measures (OR: 0.45, 95% CI: 0.21, 0.98) (Table 3). Stunting worsened in both study arms during the study period, however this decline was less in the PICHA7 arm (48% stunting at baseline to 52% at the the 12-month follow-up in the PICHA7 arm and 52% to 63% in the standard arm). There were no other significant differences in stunting, wasting, or underweight identified between study arms.

**Table 3.** Effects of PICHA7 Program on Child Growth at the 12 Month Timepoint.

| 12 Month Timepoint | Standard Message Arm |  |  |  | PICHA7 Arm |  |  |  | Effect Size or Odds Ratio (95% CI) |  |
| --- | --- | --- | --- | --- | --- | --- | --- | --- | --- | --- |
|  | Mean | SD | % | N | Mean | SD | % | N | Unadjusted | Adjusted Model<sup>1</sup> |
| **Children 0-4 years** |  |  |  |  |  |  |  |  |  |  |
| Stunting (HAZ) | 0.63 | 0.48 | 63% | 238 | 0.52 | 0.50 | 52% | 244 | 0.65 (0.43, 0.98) | 0.45 (0.21, 0.98) |
| Underweight (WAZ) | 0.25 | 0.44 | 25% | 236 | 0.21 | 0.41 | 21% | 245 | 0.81 (0.49, 1.30) | 1.11 (0.49, 2.54) |
| Wasting (WHZ) | 0.03 | 0.17 | 3% | 235 | 0.04 | 0.20 | 4% | 245 | 1.41 (0.53, 3.73) | 1.79 (0.32, 10.09) |
| **Children 0-2 years** |  |  |  |  |  |  |  |  |  |  |
| Stunting (HAZ) | 0.61 | 0.49 | 61% | 84 | 0.56 | 0.49 | 56% | 89 | 0.83 (0.44, 1.55) | 0.97 (0.31, 3.06) |
| Underweight (WAZ) | 0.17 | 0.38 | 17% | 82 | 0.27 | 0.45 | 27% | 89 | 1.79 (0.83, 3.89) | 1.17 (0.36, 3.83) |
| Wasting (WHZ) | 0.04 | 0.19 | 4% | 82 | 0.06 | 0.23 | 6% | 89 | 1.57 (0.36, 6.79) | 0.58 (0.03, 10.28) |

1. Model adjusted for baseline HAZ, WAZ, and WHZ; CI: Confidence Interval 2. Length used for children less than 2 years of age

Compared to the standard arm, handwashing with soap at stool- or food-related events was significantly higher in the PICHA7 arm at all timepoints (Week 1: OR: 11.4, 95% CI: 5.51, 26.6 to Month 12: OR: 11.8, 95% CI: 6.41, 21.7) (Table 4). This impact was similar for both stool- and food-related events individually. Relative to the WHO water quality guideline of <1 CFU/100 ml of *E.coli*, the PICHA7 arm had significantly higher water quality compared to the standard arm at all timepoints (Week 1: OR: 6.48, 95% CI: 2361, 16.1 to Month 12: OR: 4.28, 95% CI: 1.83, 10.0). Relative to the WHO free chlorine guidelines for household water treatment, PICHA7 arm households had significantly higher free chlorine >0.2 and >0.5 mg/L compared to the standard arm households at all timepoints (>0.2 mg/L free chlorine: Week 1: OR: 13.7, 95% CI: 7.03, 26.8 to Month 12: OR: 17.5, 95% CI: 8.71, 35.1) (>0.5 mg/L free chlorine: Week 1: OR: 84, 95% CI: 28.2, 250.1, Month 6: OR: 40.3, 95% CI: 13.5, 120.5).

**Table 4.** Effects of the PICHA7 Program on Individuals Handwashing with Soap at Stool and Food Related Events During 5-Hour Structured Observation (Individuals Over 2 Years) and Household Stored Drinking Water Quality and Chlorination Assessed During Unannounced Spot Checks.

| Timepoint | 1 Week |  | 1 Month |  | 3 Months |  | 6 Months |  | 9 Months |  | 12 Months |  |
| --- | --- | --- | --- | --- | --- | --- | --- | --- | --- | --- | --- | --- |
|  | % | OR (95% CI) | % | OR (95% CI) | % | OR (95% CI) | % | OR (95% CI) | % | OR (95% CI) | % | OR (95% CI) |
| **Individuals Handwashing with Soap at a Stool or Food Related Event** |  |  |  |  |  |  |  |  |  |  |  |  |
| Standard Message Arm (N=617) | 10% | - | 12% | - | 8% | - | 5% | - | 4% | - | 7% | - |
| PICHA7 Arm (N=693) | 57% | 11.4 (5.51, 26.6) | 49% | 7.09 (3.41, 14.7) | 46% | 8.34 (3.15, 22.1) | 51% | 17.3 (7.37, 40.6) | 49% | 18.5 (7.12, 48.3) | 48% | 11.8 (6.41, 21.7) |
| **Individuals Handwashing with Soap at a Food Related Event** |  |  |  |  |  |  |  |  |  |  |  |  |
| Standard Message Arm (N=564) | 8% | - | 10% | - | 8% | - | 4% | - | 4% | - | 7% | - |
| PICHA7 Arm (N=651) | 52% | 11.6 (4.92, 27.2) | 47% | 7.27 (3.28, 16.1) | 42% | 7.95 (2.81, 22.5) | 46% | 17.5 (6.62, 46.2) | 44% | 15.6 (4.98, 49.1) | 44% | 9.09 (4.62, 17.9) |
| **Individuals Handwashing with Soap at a Stool Related Event** |  |  |  |  |  |  |  |  |  |  |  |  |
| Standard Message Arm (N=411) | 6% | - | 6% | - | 7% | - | 2% | - | 3% | - | 4% | - |
| PICHA7 Arm (N=455) | 54% | 13.1 (5.04, 33.7) | 36% | 8.18 (3.35, 19.9) | 48% | 11.4 (3.02, 43.2) | 51% | 38.4 (8.80, 167.4) | 45% | 22.9 (6.43, 81.9) | 44% | 20.2 (8.14, 49.9) |
| **Households with Stored Drinking Water with <1 CFU/100 ml *E.coli*** |  |  |  |  |  |  |  |  |  |  |  |  |
| Standard Message Arm (N=134) | 61% | - | 67% | - | 57% | - | 56% | - | 61% | - | 48% | - |
| PICHA7 Arm (N=151) | 90% | 6.48 (2.61, 16.1) | 92% | 5.76 (2.08, 16.0) | 91% | 7.13 (2.20, 23.1) | 95% | 15.7 (4.12, 59.1) | 82% | 2.86 (1.04, 7.91) | 85% | 4.28 (1.83, 10.0) |
| **Households with Stored Drinking Water with Free Chlorine >0.2 mg/L** |  |  |  |  |  |  |  |  |  |  |  |  |
| Standard Message Arm (N=154) | 30% | - | 27% | - | 34% | - | 40% | - | 24% | - | 25% | - |
| PICHA7 Arm (N=161) | 86% | 13.7 (7.03, 26.8) | 75% | 8.00 (4.17, 15.4) | 82% | 8.59 (3.96, 18.7) | 77% | 5.02 (2.60, 9.69) | 84% | 16.5 (7.17, 38.1) | 85% | 17.5 (8.71, 35.1) |
| **Households with Stored Drinking Water with Free Chlorine >0.5 mg/L** |  |  |  |  |  |  |  |  |  |  |  |  |
| Standard Message Arm (N=154) | 4% | - | 2% | - | 5% | - | 5% | - | 2% | - | 1% | - |
| PICHA7 Arm (N=161) | 78% | 84.0 (28.2, 250.1) | 64% | 76.4 (17.8, 329.1) | 74% | 58.0 (16.3, 205.5) | 67% | 40.3 (13.5, 120.5) | 78% | ‡ | 76% | ‡ |

OR: Odds Ratio; CI: Confidence Interval; CFU: Colony Forming Units; Water quality analyses adjusted for water source *E.coli* concentration. ‡ not estimable due to the low prevalence of the outcome.

## Discussion

Delivery of the PICHA7 program to diarrhea patient households resulted in significantly lower diarrhea prevalence and stunting compared to those receiving the standard government of DRC ORS and basic WASH message. Severe diarrhea including diarrhea with rice water stool (key cholera symptom) and diarrhea healthcare facility visits were also halved. Furthermore, PICHA7 led to improvements in handwashing with soap and water treatment behaviors and stored drinking water quality which program households sustained for 12 months. These findings suggest that the targeted PICHA7 WASH program which combines mHealth with quarterly in-person visits can be delivered to reduce diarrhea and improve child growth in the DRC among diarrhea patient households.

The findings from the PICHA7 RCT in DRC are consistent with our previous RCT of the similar CHoBI7 WASH program in Bangladesh which found CHoBI7 sigificantly lowered diarrhea and stunting among diarrhea patient households.^22^ This promising result suggests that this targeted WASH program for diarrhea patients households delivered through mHealth and quarterly in-person visits has the potential to reduce diarrhea and improve child growth in two distinct contexts. Our study site in urban DRC differs from our site in urban Bangladesh because of a lower proportion of safely managed water services (24% DRC vs. 54% Bangladesh)^23^, lower improved sanitation (53% vs. 90%)^23^, lower population density (13,449 (Bukavu) individuals per sq km vs. 41,000 (Dhaka))^24^, higher prevalence of moderate/severe food insecurity (77% vs. 31%)^25^, and higher household size (6.5 individuals per household (from our PICHA7 RCT) vs. 3.4^22^). Our formative research was critical to tailor the PICHA7 program to this different context.^11^

An important finding from our PICHA7 and CHoBI7 RCTs is that WASH behavior change is possible without needing frequent promoter home visits. In CHoBI7 only a single in-person visit in a healthcare facility and two home visits was delivered^22^ and in PICHA7 an in-person visit in the healthcare facility followed by quarterly in-person home visits was delivered for 12 months. This is in contrast to previous studies where almost all that reported a positive effect on diarrhea had biweekly or more frequent visits.^26^ Community health workers currently conduct household visits in our study area in the DRC, therefore PICHA7 delivery quarterly with chlorine tablet distribution has the potential to be integrated into this existing community health worker programming. Furthermore, mHealth appears to be a promising approach to reinforce WASH recommendations delivered during in-person visits to facilitate WASH behavior change. The process evaluation of the PICHA7 mHealth program found that 84% of text messages were received and 90% of voice calls were answered during the 12-month program (Sanvura et al. submitted). However, PICHA7 and CHoBI7 are the only RCTs of WASH mHealth programs. Additional trials are needed in rural settings and other settings globally to evaluate the impact of WASH mHealth programs on diarrheal diseases and child growth.

The effect of the PICHA7 program on diarrhea prevalence was stronger than the effect we observed in the CHoBI7 RCT over the same 12-month program period (diarrhea reduction for all ages: 61% PICHA7 vs. 29% CHoBI7). This may be because of the higher diarrhea prevalence for children 0-4 years in the PICHA7 RCT (32% PICHA7 vs. 21% CHoBI7) allowing for more room for WASH delivery improvement. Additionally, this may be because of the higher intensity of the PICHA7 program which had quarterly in-person intervention visits over a 12 month period compared to only two in-person visits during the 7-day high risk period after diarrhea patient admission in the CHoBI7 program.

The PICHA7 program was found to be effective in reducing severe diarrhea by half, including diarrhea with rice water stool (cholera symptom) and diarrhea healthcare facility visits. Our findings are consistent with a study conducted in DRC which found hygiene kit delivery to suspected cholera patient households resulted in a 56% lower incidence of suspected cholera (defined by diarrhea, vomiting, or a diarrhea healthcare facility visit).^27^ These findings are also consistent with our CHoBI7 RCT in Bangladesh which found that delivery of the CHoBI7 program to cholera patient households (bacterial culture confirmed) resulted in a 47% reduction in cholera infections confirmed by bacterial culture among household members.^28^ The findings from the PICHA7 and CHoBI7 RCTs suggest that this type of targeted WASH for diarrhea patient households may be effective in reducing severe forms of diarrhea such as cholera.

The PICHA7 program sigificantly reduced stunting among children 0-4 years. This finding is consistent with a RCT in Mali which found a WASH program focused on community-led-total-sanitation (CLTS) reduced stunting.^29^ The RCTs of PICHA7 in DRC, CHoBI7 in Bangladesh, and CLTS in Mali are the only three RCTs to date, to our knowledge, that have found a sigificant association between WASH delivery and improvements in children’s linear growth. In the CLTS RCT in Mali stunting was reduced, but not diarrhea^29^, whereas the PICHA7 and CHoBI7 RCTs are the only WASH RCTs that sigificantly reduced both stunting and diarrhea among young children.^22^ For the interpretation of PICHA7 RCT stunting findings it should be noted that stunting worsened in both study arms during the 12-month study period (48% to 52% in the PICHA7 arm and 52% to 63% in the standard arm), however less for those participants in the PICHA7 arm. We attribute the impact of the PICHA7 program on stunting to the large reduction in diarrhea prevalence (>60% reduction for children <5 years), the high adherence observed for handwashing with soap and water treatment, and high baseline stunting prevalence (50%) which likely allowed more room for improvement with WASH intervention delivery.

No sigificant impact of PICHA7 was observed for any growth parameter other than stunting. One potential explanation for this findings is the need for PICHA7 to be combined with food supplements for young children. Our previous work at this study site found that only 26% of children consumed five or more minimum dietary diversity food groups^30^, and moderate/severe food insecurity in the DRC is estimated to be 77%.^25^ However, previous trials investigating the impact of combined WASH and nurition program did not observe an impact on children growth.^31,32^ Future RCTs are needed to evaluate if combining WASH interventions similar to PICHA7 and CHoBI7 with nutritional supplements can facilitate improvements in wasting and other growth parameters in children.

This study has limitations. First, we relied on self- or caregiver-reported diarrhea and did not assess the presence of enteric pathogens among study participants to determine the etiology of diarrhea. Future studies will investigate the impact of PICHA7 on specific enteric pathogens. Second, we only assessed the impact of PICHA7 among diarrhea patient households. This group was selected since they represent a high-risk population for diarrheal diseases, however studies have shown that other households residing within 20 to 30 meters to diarrhea patients households are also at higher risk of diarrheal diseases.^9,10^ Future studies should evaluate the impact of delivery of PICHA7 in a ring around diarrhea patients on reducing diarrhea in these diarrheal disease hotspots. Additionally, because all PICHA7 intervention components were combined and not evaluated separately it is not possible to determine which PICHA7 intervention component was most influential in the success of this program. Future studies should investigate the efficacy of the individual components of PICHA7 on diarrheal disease and child growth.

The PICHA7 RCT has several strengths. First is the monthly clinical surveillance data for both children and adults, building on previous WASH studies that focused mostly on diarrhea among children 0-4 years. Second is the inclusion of severe diarrhea measures including diarrhea with rice water stool (symptom of cholera), and healthcare facility visits for diarrhea. Most WASH studies focus on mild to moderate diarrhea. Third, the inclusion of 115 healthcare facilities in the PICHA7 RCT patient recruitment which allowed for the intervention to be tested in wide range of public and private health facilities.

## Conclusion

The PICHA7 WASH program significantly lowered diarrhea and stunting and led to sustained WASH behavior during the 12-month program period. Furthermore, PICHA7 resulted in lower severe diarrhea including diarrhea with rice water stool and healthcare facility visits for diarrhea. We are currently partnering with the DRC Ministry of Health to develop strategies to scale PICHA7 as part of the DRC National Cholera Control Plan. Our findings suggest that the PICHA7 program which combines mHealth and quarterly in-person visits presents a promising approach to reduce diarrhea and improve child growth for diarrhea patient households in the DRC. Future trials are needed in diverse settings globally to investigate the health impacts of WASH programs combining mHealth and in-person visits for intervention delivery.

## Data Availability

All data produced in the present study are available upon reasonable request to the authors.

## Acknowledgements

We thank the study participants and the following individuals for their support with this study: Dr. Placido Welo, Dr. Jean Claude Kulondwa, and Dr. Roger Boketsu. We thank all DRC Ministry of Health in South Kivu staff.

## Abbreviations

PICHA7: Preventative-Intervention-for-Cholera-for-7-Days
DRC: Democratic Republic of the Congo
mHealth: mobile health
PR: Prevalence Ratio
OR: Odds Ratio
CI: Confidence Interval
WASH: water, sanitation, and hygiene
RCT: randomized controlled trial
HAZ: height-for-age
WAZ: weight-for-age
WHZ: weight-for-height
WHO: World Health Organization
CFU: colony forming units
CHoBI7: Cholera Hospital Based Intervention for 7 Days

