## Supplementary File 1 for "Effects of a Water, Sanitation, and Hygiene Program on Diarrhea and Child Growth in the Democratic Republic of the Congo: A Cluster-Randomized Controlled Trial of the Preventative-Intervention-for-Cholera-for-7-Days (PICHA7) Program"

**Supplementary Appendix 1**

The PICHA7 program is initially delivered during a health facility visit by a health promoter bedside to a diarrhea patient and their accompanying household members during the time of illness. The promoter delivers a 30-minute session which includes a pictorial WASH module and videos testimonials (shown on a mobile phone) from diarrhea patients on how they have benefited from the PICHA7 program. A diarrhea prevention package is also provided containing the following: 32 chlorine tablets for water treatment ((Aquatabs [67 mg sodium dichloroisocyanurate]; Medentech, Wexford, Ireland), a soapy water bottle containing water and detergent powder, a handwashing station, and a water vessel with a lid and tap to ensure safe water storage. After health facility delivery of the program, diarrhea patient households receive weekly voice, IVR, and text messages to their mobile phone from the PICHA7 mHealth program over a 12-month period. These mobile messages are sent using the EngageSPARK platform. Two characters deliver the PICHA7 mHealth messages: “Dr. Picha”, a doctor at a cholera treatment center who treats cholera and severe diarrhea patients, and “Mwanza”, a mother of a young child that fell ill with cholera and needed to go a cholera treatment center for treatment. Two 30-minute home visits are conducted by a promoter during the 7-day high risk period for diarrheal disease in the diarrhea patient household. The first visit is conducted the day after enrollment of the diarrhea patient and the second visit at Day-7 after enrollment. This is followed by 15-minute quarterly in-person home visits (every 3 months) by a health promoter to provide 32 additional chlorine tablets and reinforce the promoted WASH behaviors from the earlier in-person visits.
