## Supplementary figures and images for "Effects of a Water, Sanitation, and Hygiene Program on Diarrhea and Child Growth in the Democratic Republic of the Congo: A Cluster-Randomized Controlled Trial of the Preventative-Intervention-for-Cholera-for-7-Days (PICHA7) Program"

### Supplementary File 2

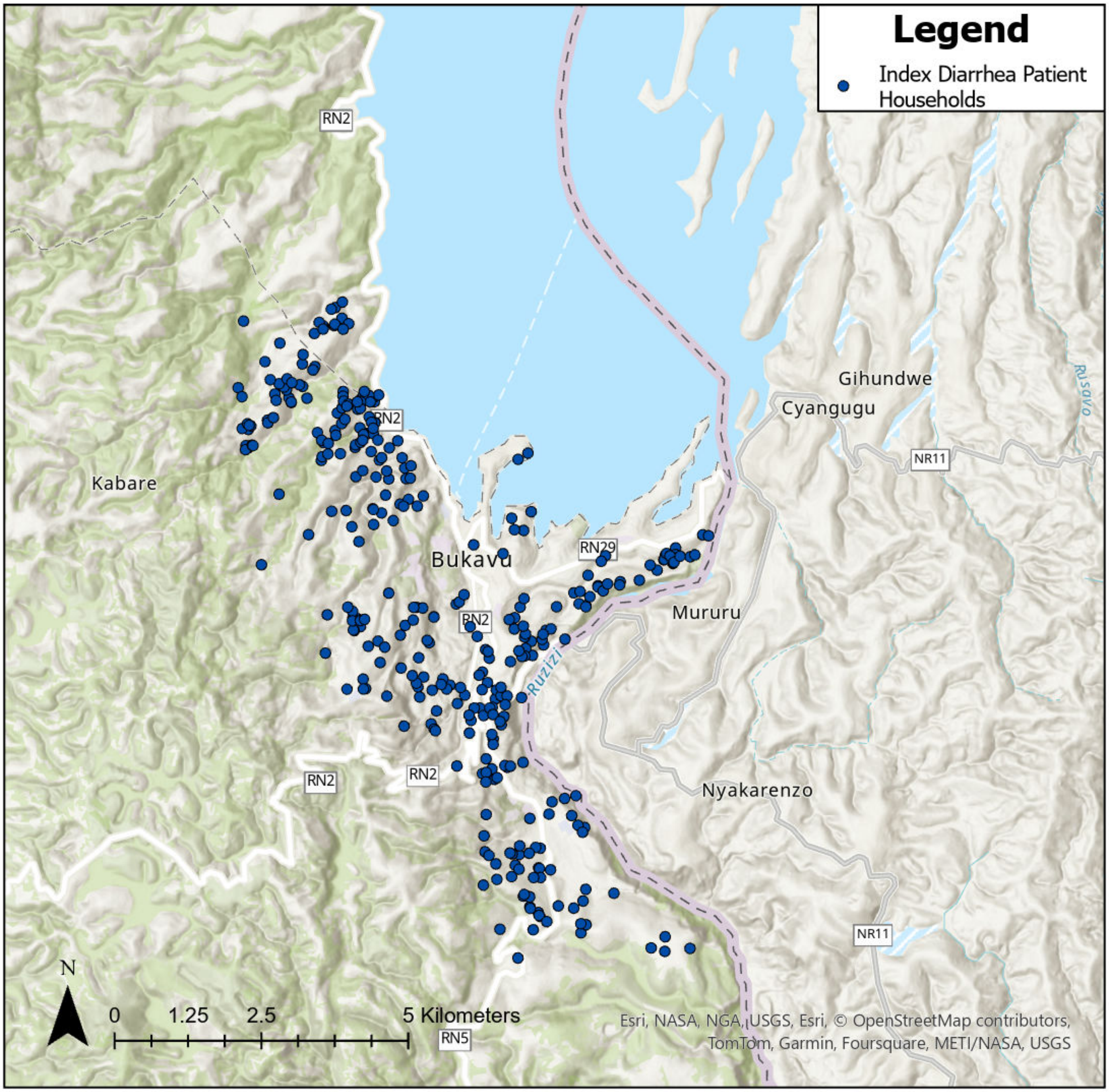
